# GPT-4 Improves Readability of Institutional Heart Failure Patient Education Materials: An Observational Study

**DOI:** 10.1101/2024.11.08.24316997

**Authors:** Ryan C. King, Jamil S. Samaan, Joseph Haquang, Vishnu Bharani, Samuel Margolis, Nitin Srinivasan, Yuxin Peng, Yee Hui Yeo, Roxana Ghashghaei

## Abstract

**Introduction:** Heart failure management involves comprehensive lifestyle modifications such as daily weights, fluid and sodium restriction, and blood pressure monitoring placing additional responsibility on patients and caregivers with successful adherence often requiring extensive counseling and understandable patient education materials (PEMs). Prior research has shown PEMs related to cardiovascular disease often exceed the American Medical Association’s 5_th_-6_th_ grade recommended reading level. The large language model (LLM) Chat Generative Pre-trained Transformer (ChatGPT) may be a useful tool for improving PEM readability.

**Materials and Methods:** A total of 143 heart failure PEMs were collected from the websites of the top 10 institutions listed on the 2022-2023 US News & World Report for “Best Hospitals for Cardiology, Heart & Vascular Surgery”. PEMs were individually entered into GPT-4 (Version updated 20 July 2023) preceded by the prompt “please explain the following in simpler terms”. The readability of the institutional PEM and ChatGPT revised PEM were both assessed using *Textstat* library in Python and the *Textstat readability* package in R software. The accuracy and comprehensiveness of revised GPT-4 PEMs were assessed by a board-certified cardiologist.

**Results:** The Flesch-Kincaid grade reading level ranged from 8th grade to college freshman with a median of 10th grade vs 6^th^ to 8^th^ grade with a median of 7^th^ grade for institutional PEMs and GPT-4 PEMs (p< 0.001), respectively. There were 13/143 (9.1%) institutional PEMs below the 6_th_ grade reading level which improved to 33/143 (23.1%) after revision by GPT-4 (p<0.001). No GPT-4 revised PEMs were graded as less accurate or less comprehensive compared to institutional PEMs. A total of 33/143 (23.1%) GPT-4 PEMs were graded as more comprehensive.

**Conclusions:** GPT-4 significantly improved the readability of institutional heart failure PEMs. The model may be a promising adjunct resource in addition to care provided by a licensed healthcare professional for patients living with heart failure. Further rigorous testing and validation is needed to investigate its safety, efficacy and impact on patient health literacy.

## Introduction

Heart failure affects approximately 1-2% of adults globally, with an estimated prevalence of 64 million people [1]. Treatment involves extensive patient adherence to lifestyle modifications such as daily weights, fluid and sodium restriction, and rigorous guideline-directed medication regimens. Altogether, these interventions attempt to prevent disease progression and hospital admissions, which drive most of the financial burden ($39.2-$60 billion) related to the disease [2]. Due to the complex degree of self-management required by heart failure patients, improving patient education and health literacy may play a crucial role in improving outcomes [3,4].

In the United States, the average adult’s reading comprehension level is approximately 7th-8th grade proficiency [5], resulting in the American Medical Association (AMA) recommendation of written patient education materials (PEMs) being at a 5th-6th grade reading level [6]. However, a 2019 readability analysis of cardiovascular-disease-related PEMs reported that the mean reading level of materials was 10.5, comparable to that of a high school sophomore [7]. Inadequate health literacy has been associated with increased relative risk of emergency department visits, hospitalizations, and mortality for heart failure patients [4,8], highlighting the need for accessible, readable, and high-quality PEMs.

Chat Generative Pre-trained Transformer (ChatGPT) is a Large Language Model (LLM) that is gaining widespread public adoption [9]. With an increasing number of patients seeking health information online [10], the model has the potential to enhance patient health education and address the complexity of heart failure PEMs. As ChatGPT’s acceptance and usage have increased, initial research involved evaluating the model’s accuracy and reliability. Several studies have shown that ChatGPT provides appropriate, accurate, and reliable knowledge across a wide range of cardiac and non-cardiac medical conditions, including heart failure [11–16]. In addition to accuracy, ChatGPT has been found to deliver more empathetic responses to real world patient questions than physicians in online forums [17]. As prior data regarding accuracy has been promising, an emerging focus has been on investigating the readability of the model’s output.

Prior studies have shown ChatGPT provides accurate and comprehensive responses to questions related to heart failure, while another demonstrated its responses were at a college reading level highlighting the need for further assessment of the readability of GPT’s outputs [12,18]. Similarly, another study examining GPT-4’s responses related to amyloidosis showed while responses were often accurate and comprehensive, the average readability of responses ranged from 10.3 (high school sophomore) to 21.7 (beyond graduate school) grade level [16]. We aimed to expand on the previous literature by assessing the readability of heart failure-related online PEMs from renowned cardiology institutions, assessing GPT-4’s ability to improve the readability of these PEMs, and compare the accuracy and comprehensiveness between institutional PEMs and GPT-4’s revised PEMs.

## Methods

### Institutional Patient Education Materials

There were 143 PEMs (Suppl. file 1) related to heart failure collected in July 2023 from the top 10 ranked cardiology institutions (deidentified) listed on the 2022-2023 US News & World Report website as “Best Hospitals for Cardiology, Heart & Vascular Surgery”. These PEMs include frequently asked questions (FAQs) presented as text descriptions of various aspects of heart failure such as causes, symptoms, medications, and procedures. Duplicate institutional PEMs were included given education materials varied between institutions and readability of each PEM is the primary outcome of interest.

### GPT-4 Response Generation

Each institution’s PEMs were entered into GPT-4 (Version updated 20 July 2023) preceded by the prompt “Please explain the following in simpler terms.” GPT-4 was accessed using the OpenAI website interface. Default model settings were used (temperature, max tokens, etc.). The “new chat” function was used for each PEM thus creating a new conversation without record of prior inputs. Materials containing non-text components (images or videos) were excluded.

### Readability Assessment

The readability of institutional PEMs and GPT-4 revised PEMs were then assessed using the following validated formulas: Flesch Reading Ease score (FRE) [19], Flesch-Kincaid Grade Level (FKGL) [20], Gunning Fog Index (GFI) [21], Coleman-Liau Index (CLI) [22], Simple Measure of Gobbledygook (SMOG) Index [23], Automated Readability Index (ARI) [24]. The FRE score, measured on a scale of 0 to 100, indicates a text with a higher score has better ease of understanding. The remaining formulas directly translate a score into its corresponding U.S. reading grade level such as a score of 10 meaning a 10th grade reading level. These metrics derive their scores from the mean length of sentences and words used in a given text. In contrast to the FRE, the other formulas yielding a lower score corresponds to an easier level of understanding. The readability formulas were assessed using the *Textstat* library in Python (Python Software Foundation) and the *Textstat readability* package in R software (R Foundation for Statistical Computing).

### Accuracy and Comprehensiveness

Accuracy and comprehensiveness of GPT-4’s revised PEMs (Suppl. file 1) were assessed by an actively practicing board-certified cardiologist at a tertiary academic medical center. During grading, the reviewer was not blinded to the source of responses. The reviewer used the following grading scale when grading the original institutional PEMs and revised GPT-4 PEM.

“Compared to the institutional PEM the GPT-4 revised PEM is”:

1. Less accurate
2. Equally accurate
3. More accurate

“Compared to the institutional PEM, the GPT-4 revised PEM is”:

1. Less comprehensive
2. Equally comprehensiveness
3. More comprehensive

### Statistical Analysis

Descriptive statistics are presented as medians and interquartile ranges (IQR). Readability metrics for institutional PEMs and GPT-4 revised PEMs were compared using the Mann-Whitney U test. Further sub-analysis was performed investigating the proportion of PEMs meeting the 6^th^ reading grade level recommendation by the AMA among institutional and GPT-4 revised PEMs. Statistical analysis was conducted using IBM SPSS version 2

**Figure 1.**
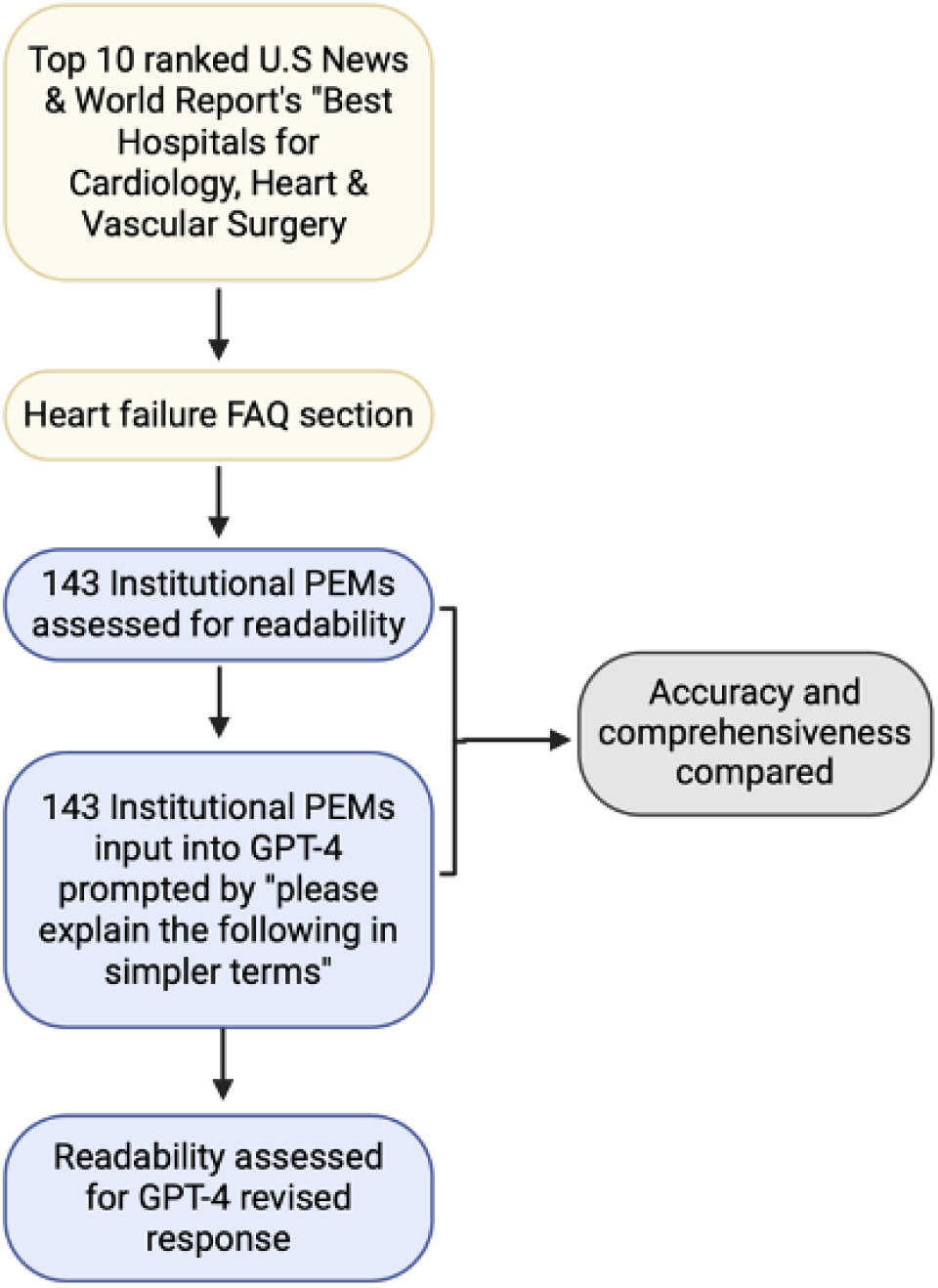
Diagram of institutional heart failure patient education materials curation, GPT-4 revised patient education materials generation and subsequent assessment of readability, accuracy, and comprehensiveness. Figure created using biorender.com.

**Figure 2.**
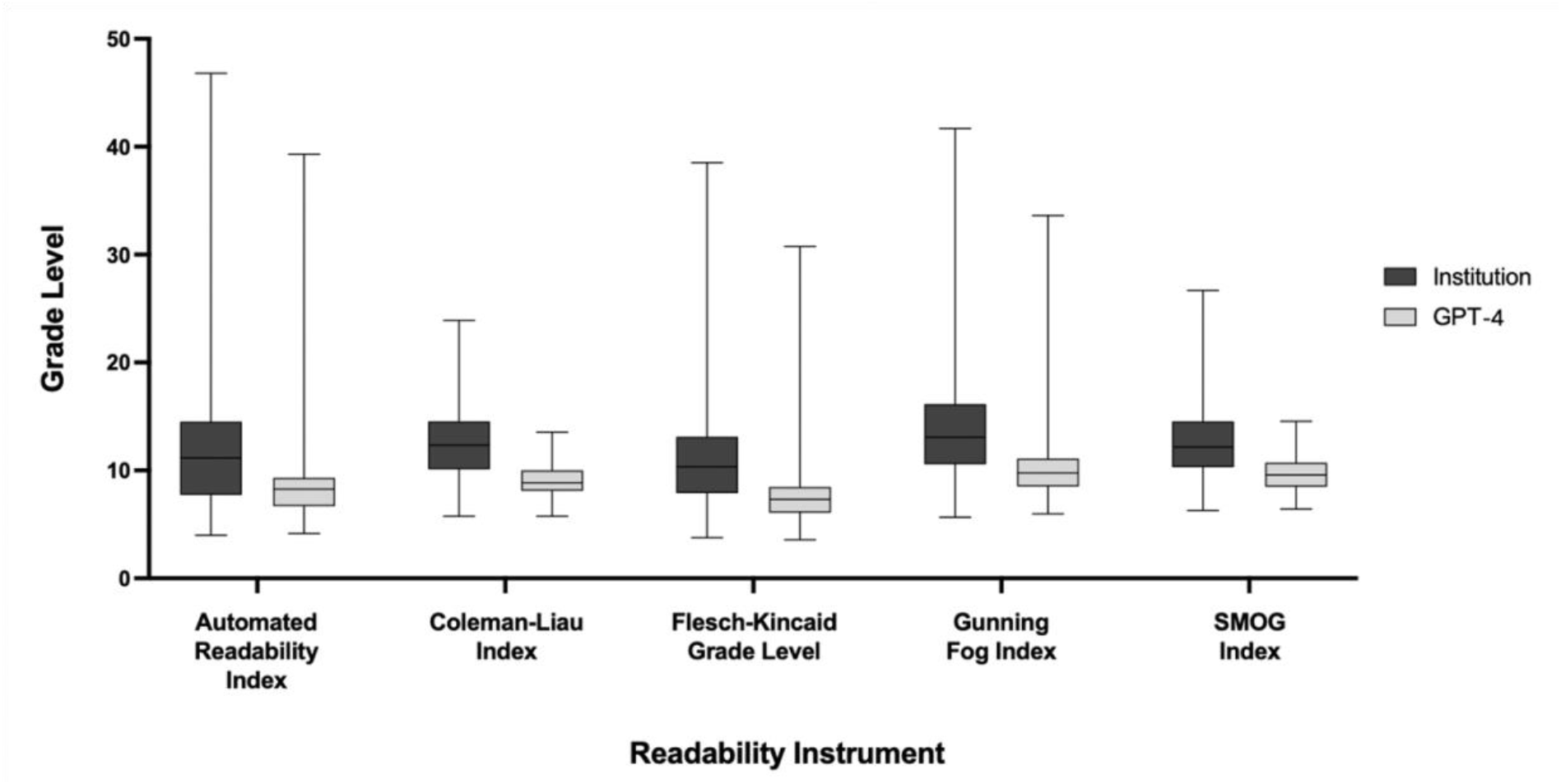
Box and whiskers plot of median readability scores across five metrics including Automated Readability Index, Coleman-Liau Index, Flesch-Kincaid Grade Level, Gunning Fog Index, SMOG Index for institutional and GPT-4 revised PEMs. **GPT-4:** Generative Pre-trained Transformer-4; **PEMs:** Patient education materials.

**Table 1.** Comparison of readability of institutional and GPT-4 revised PEMs. **SMOG**: Simple Measure of Gobbledygook; **IQR**: Interquartile Range; **GPT-4**: Generative Pre-trained Transformer-4.

|  | Institutional |  |  | GPT-4 |  |  | p-value |
| --- | --- | --- | --- | --- | --- | --- | --- |
| Readability Metrics | Median | IQR (25th, 75th) |  | Median | IQR (25th, 75th) |  |  |
| Flesch Reading Ease score | 48.6 | 38.0 | 63.3 | 72.2 | 66.2 | 77.5 | <.001 |
| Flesch-Kincaid Grade Level | 10.3 | 7.9 | 13.1 | 7.3 | 6.1 | 8.5 | <.001 |
| Gunning Fog Index | 13.1 | 10.6 | 16.2 | 9.8 | 8.5 | 11.1 | <.001 |
| Coleman-Liau index | 12.3 | 10.1 | 14.5 | 8.9 | 8.1 | 10.0 | <.001 |
| SMOG index | 12.2 | 10.3 | 14.6 | 9.6 | 8.5 | 10.7 | <.001 |
| Automated readability index | 11.2 | 7.7 | 14.5 | 8.3 | 6.7 | 9.3 | <.001 |

**Table 2.** Comparison of the proportion of PEMs meeting the AMA’s recommended 5^th^-6^th^ grade reading level between institutional and GPT-4. **PEM**: Patient education materials; **GPT-4**: Generative Pre-trained Transformer-4; **AMA**: American Medical Association.

|  | **<=6 Reading Level** | **>=6 Reading Level** | **Percent Meeting**<br>**AMA**<br>**Recommendation** |
| --- | --- | --- | --- |
| **Institutional Flesch-Kincaid Grade Level** | 13.0 | 130.0 | 9.1% |
| **GPT Flesch-Kincaid Grade Level** | 33.0 | 110.0 | 23.1% |

**Table 3.**
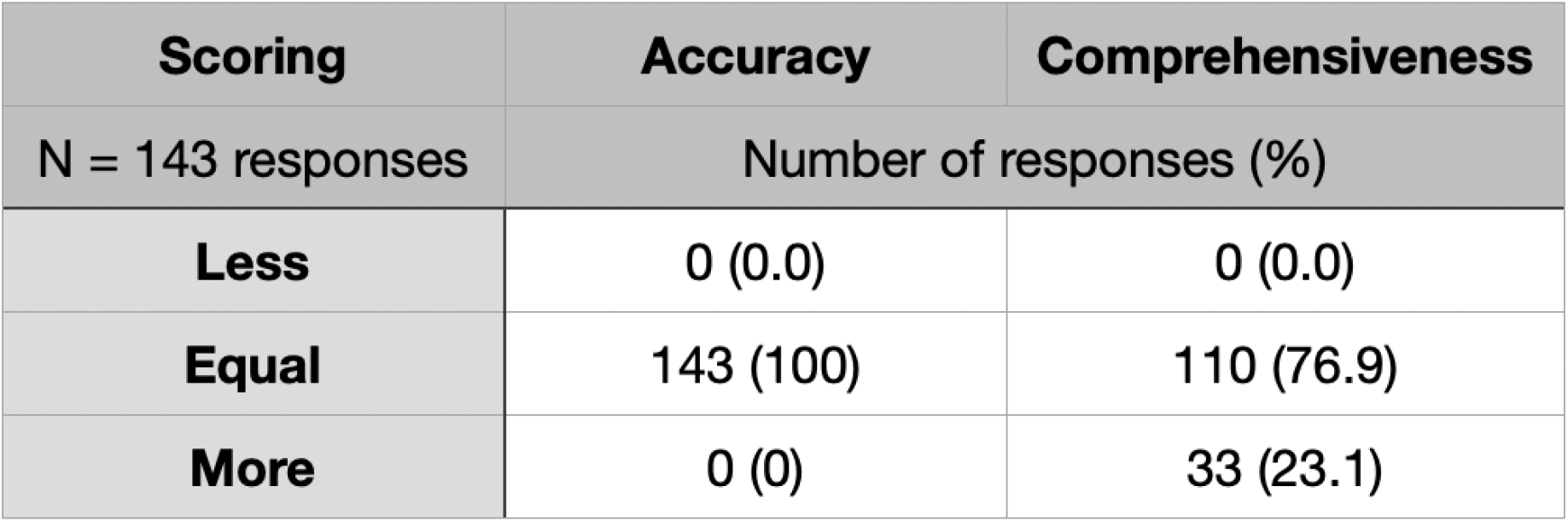
Evaluation of GPT-4’s accuracy and comprehensiveness of revised PEMs compared to institutional PEMs. **PEM**: Patient education materials; **GPT-4**: Generative Pre-trained Transformer-4.

| **Scoring** | **Accuracy** | **Comprehensiveness** |
| --- | --- | --- |
| N = 143 responses | Number of responses (%) |  |
| **Less** | 0 (0.0) | 0 (0.0) |
| **Equal** | 143 (100) | 110 (76.9) |
| **More** | 0 (0) | 33 (23.1) |

## Results

### Readability Assessment

Readability analysis revealed GPT 4’s revised PEMs were significantly more readable compared to institutional PEMs across all six metrics (p<.001). The Flesch Reading Ease score increased from a median institutional score of 48.6 (IQR: 38.0, 63.3; p<.001) to 72.2 (IQR: 66.2, 77.5; p<.001) after GPT-4 revision. The Flesch-Kincaid Grade level also saw improvement, decreasing from an institutional median reading level of 10^th^ grade (IQR: 7.9,13.1; p<.001) to 7^th^ grade (IQR: 6.1, 8.5; p<.001) after GPT-4 revision. Furthermore, the institutional Automated Readability Index of 11.2 (IQR: 7.7, 14.5; p<.001) improved to 8.3 (IQR: 6.7, 9.3; p<.001) after GPT-4 revision. The other readability metrics (Gunning Fog, Coleman-Liau, and SMOG) also showed improved scores after GPT revision: 9.8 (IQR: 8.5, 11.1; p<.001), 8.9 (IQR: 8.1, 10.0; p<.001), and 9.6 (IQR: 8.5, 10.7; p<.001), respectively, compared to the median institutional scores of 13.1 (IQR: 10.6, 16.2), 12.3 (IQR: 10.1, 14.5), and 12.2 (IQR: 10.3, 14.6). Before GPT-4 revision, 9.1% (13/143) of institutional PEMs met the AMA’s recommended 6th-grade reading level. However, after GPT-4’s revision, 23.1% (33/143) of PEMs met the 6^th^-grade recommendation. On average, GPT-4 revision led to a 3.6 reading grade level reduction.

An example of this simplification in reading level was seen when describing different types of heart failure. The institutional PEM described right sided heart failure most often resulting from left sided heart failure due to increased pressure from the left ventricle not propelling blood to the rest of the body. However, GPT-4 provided a more basic explanation using an analogy of ventricles being small rooms and gave a more simplified explanation of right sided heart failure as a result of left sided heart failure. In another example, when explaining the various causes of heart failure, one institutional PEM provided a list of etiologies such as “heart valve disease” or “coronary artery disease” without a description compared to GPT-4 which more thoroughly described the role of each cause in relation to heart failure in simple language.

### Accuracy and Comprehensiveness

Following review by a board-certified cardiologist, 33/143 GPT-4 PEMs (23.1%) were graded as more comprehensive than the corresponding institutional PEMs. Additionally, all 143/143 (100%) GPT-4 PEMs were graded as equally accurate as their institutional PEM counterpart.

## Discussion

### Principal Results

LLMs are a rapidly developing technology with the potential to enhance the delivery of patient education materials (PEMs) to patients of all levels of health literacy. In this study, we expanded on existing research that evaluated ChatGPT’s ability to generate accurate and reliable answers to heart failure questions by examining GPT-4’s ability to improve the readability of institutional PEMs. Our analysis shows GPT-4, when prompted, was able to significantly enhance the readability of institutional PEMs for common patient heart failure questions. After evaluation by a board-certified cardiologist, all GPT-4’s revised PEMs were graded as equally accurate and many graded as more comprehensive as institutional PEMs, with no revised PEMs graded as less accurate or less comprehensive. GPT-4’s capabilities to provide accurate, comprehensive and readable PEMs in real-time and in a conversational manner underscores the future potential of LLMs to enhance patient education and ultimately patient health literacy.

### Comparison with Prior Work

Previous research has demonstrated that ChatGPT possesses a broad knowledge base comprising various medical conditions, including cirrhosis, hepatocellular carcinoma, and bariatric surgery [14,15,25,26]. Its knowledge base also spans cardiovascular diseases such as acute coronary syndrome [11,27], heart failure [12], atrial fibrillation [28], and even rare disorders like amyloidosis [16]—a multi-systemic infiltrative disease. Specifically, regarding amyloidosis, while GPT-4 provided accurate, comprehensive, and reliable answers to gastrointestinal, neurologic, and cardiology queries, the average Fleisch-Kinkaid reading grade level of responses was 15.5 (college level), significantly exceeding the recommended 6^th^ grade reading level set forth by the AMA [16]. Similar results were shown when examining responses to the surgical treatment of retinal diseases and hypothyroidism in pregnancy [29,30].

A previous study examined ChatGPT’s ability to simplify the readability of responses to bariatric surgery frequently asked questions [31]. GPT-4 reduced the average grade reading level of PEMs from 11^th^ (high school junior) to 6^th^ grade, aligning with the AMA’s recommendation. Another study also showed GPT-4 improved the readability of cardiovascular magnetic resonance reports, reducing the average reading level from 10th grade to 5th grade while maintaining high factual accuracy [32]. Our study further contributes to this body of work by demonstrating GPT-4’s ability to improve the median readability of institutional PEMs from 10.3 (high school sophomore) to 7.3 (7^th^ grade) while maintaining accuracy and often enhancing comprehensiveness. The enhanced readability, achieved without specific instructions to simplify to a particular education level, underscores the potential of ChatGPT in fostering better patient understanding of heart failure-related information.

### Limitations, and Ethical Considerations

ChatGPT, while adept at generating conversational answers, has inherent limitations in accuracy and privacy. The model cannot access real-time patient records, often does not cite peer-reviewed articles, or reference updated guidelines, which is crucial for accurate and evidence-based responses. Additionally, the current model may not reliably understand nuanced medical topics or accurately interpret complex medical questions [33], leading to potential patient misunderstandings. In some cases, ChatGPT may also generate answers that initially seem factual due to its confident appearing language but disseminate inaccurate information, known as artificial hallucinations [34]. Utilizing Artificial Intelligence (AI) models like ChatGPT in healthcare settings may also not guarantee secure handling of patient information as the model may collect users’ conversation data for future training. Although OpenAI does have a privacy setting allowing for disabling user data collection, prioritizing patient confidentiality will be an important aspect of development if the technology is to be used as an adjunct healthcare tool [35].

Furthermore, ChatGPT may also perpetuate social disparities due to implicit biases and contribute to accessibility gaps. Recent studies revealed that GPT-4 tended to promote outdated race-based medicine, overrepresent, or underrepresent certain racial groups and sexes depending on the circumstance and thus potentially reinforce stereotypes [36,37]. Another concern is equitable access, as patients with lower socioeconomic status often have less access to certain technology such as the internet and may have barriers to utilizing these new AI tools [38]. Altogether, these validity and ethical considerations emphasize that clinical oversight such as FDA regulation is warranted prior to LLMs incorporation in patient care [39]. This would allow for consistent monitoring of this rapidly evolving technology ensuring optimization of safety protocols with each new update of the model.

Our study has several limitations. Although we employed validated readability scoring systems as a surrogate for patient understanding, these formulas have their limitations as previously reported [40,41]. These formulas often generate a reading level score that inherently grades longer words and sentences as being more complex but are unable to assess a text’s content for structure and clarity. Our study also did not involve patients, which is essential for the comprehensive assessment of ChatGPT as a patient educational resource. Future studies would benefit from involving patients to ensure relevance of questions, preference in language used and assessment of patient understanding. A baseline assessment of a patient’s understanding of the given topic would also be beneficial to assess if ChatGPT can improve comprehension rather than relying on scoring tools. Additionally, we employed only one expert reviewer to assess the accuracy and comprehensiveness of ChatGPT’s responses. To limit the potential for bias through subjective review and promote diverse perspectives, future research would benefit from involving multiple reviewers from different backgrounds and training institutions. Our reviewer was also not blinded to the source of each PEM allowing for possible bias when evaluating accuracy and comprehensiveness. Our study could also not incorporate or interpret questions containing multimedia at the time of data collection, but with the release of multimodal LLMs, like GPT-4v, including visual aids would be another valuable component of PEMs to investigate. The PEMs used are not comprehensive of all questions that may be asked by patients which limits the generalizability of our results. Future studies utilizing real world patients and questions would be helpful to further understand the broad spectrum of questions patients may ask.

We opted for a pragmatic approach in designing the GPT-4 prompt used to revise institutional PEMs. Our focus was on ensuring the prompt reflected a simple, intuitive command that patients would be likely to use in real-world scenarios. However, exploring more intricate prompts could yield even more impressive outputs and functionality. We advocate further research into prompt engineering to better replicate natural conversations and offer specific instructions for generating higher-quality responses.

## Conclusions

Our study demonstrates GPT-4’s ability to improve the readability of institutional heart failure PEMs while also maintaining accuracy and comprehensiveness. Our results underscore the potential future utility of LLMs in improving the delivery of easy to understand and readable PEMs to patients of all health literacy levels. While ChatGPT may potentially be a valuable future tool in patient care, it should be used as a supplement to, rather than a replacement for, human expertise and judgment of a licensed healthcare professional. We recommend the development of future studies examining optimization of readability outputs, personalization, and real-world implementation.

## Acknowledgements

ChatGPT-4, version updated 16 May 2024, by OpenAI, 2023 was used in the final editing process of this manuscript to improve readability.

## Data Availability

All data produced in the present study are available upon reasonable request to the authors

## Abbreviations

AMA: American Medical Association
ARI: Automated Readability index
ChatGPT: Chat Generative Pre-trained Transformer
CLI: Coleman-Liau Index
FKGL: Flesch-Kincaid Grade Level
FRE: Flesch Reading Ease score
GFI: Gunning Fog Index
LLM: Large language model
PEMs: Patient education materials
SMOG: Simple Measure of Gobbledygook

